# The cost of the plunge: The impact and cost of a cessation of PEPFAR-supported services in South Africa

**DOI:** 10.1101/2025.04.22.25326207

**Authors:** Gesine Meyer-Rath, Lise Jamieson, Edinah Mudimu, Jeffrey W. Imai-Eaton, Leigh F. Johnson

## Abstract

**Background:** Globally, shifts in United States foreign aid policy have put HIV funding under duress. South Africa is unique in the region because its HIV programme is largely domestically funded, although donor funds and partnerships support key components. We estimated the epidemiological impact of ceased PEPFAR support for specific services and the costs and cost-effectiveness of the South African government (SAG) potentially taking over these services.

**Methods:** We used a costed version of Thembisa, a South African HIV transmission model, to simulate four scenarios: a minimum scenario assuming intervention coverage reducing proportional to PEPFAR’s funding share of specific activities in 2023; a maximum scenario assuming additional health system impacts; and sub-scenarios either with 3-year recovery (2029-2031) or no recovery to previous coverage. HIV programme costs were estimated from the provider perspective (SAG) in 2024/25 US dollars.

**Results:** Over 2025-2028, discontinuing PEPFAR funding in South Africa without replacement by SAG would result in 150,000-296,000 additional new HIV infections (29-56% increase) and 56,000-65,000 additional AIDS-related deaths (33-38%). Permanent discontinuation of currently PEPFAR-supported services over the next 20 years increases this to 1.1-2.1 million additional new HIV infections and 519,000-712,000 additional AIDS deaths. To sustain services, SAG needs to spend an additional $710 million to $1.5 billion between 2025 and 2028. Under a reduced budget, the most cost-effective interventions to preserve are ART and PrEP.

**Conclusion:** Unmanaged PEPFAR exit from South Africa threatens to undo a decade of progress towards ending AIDS as a public health threat unless services are taken over by other funders, including SAG.

## INTRODUCTION

Funding of HIV programmes in many low- and middle-income countries globally is under duress as a result of a shift in United States (US) policy regarding foreign aid. South Africa, home to the world’s largest number of people living with HIV (PLHIV), is in a unique situation. Since the start of the public-sector HIV programme 21 years ago, the majority of it has been funded from domestic resources, unlike most other high HIV prevalence countries where donor funds from the US President’s Emergency Plan for AIDS Relief (PEPFAR) and the Global Fund enable most of the response. In 2023, the last year for which complete expenditure data are available, PEPFAR accounted for 21% of South Africa’s total HIV expenditure of $1.86 billion and the Global Fund for another 3% [1]. However, this expenditure was unequally distributed across programme areas; for example, while the South African government funded 86% of antiretroviral treatment (ART) in 2023, PEPFAR funded 50% of prevention services, including substantially the voluntary male medical circumcision (VMMC) and pre-exposure prophylaxis (PrEP) programmes. Additionally, PEPFAR supported substantial treatment retention activities by community-based healthcare workers, interventions for key populations, and health system strengthening, including investments and technical assistance in logistics, supply chain management, and data systems (see Supplementary Table 1).

In late January 2025, following the executive order mandating reviews of all US foreign assistance programmes, we were tasked by the South African National Department of Health to calculate the epidemiological impact of a complete cessation of activities receiving PEPFAR support and the costs to the South African government (SAG) of taking over services paid for by PEPFAR. Shortly thereafter, a further order halted all foreign aid or assistance to South Africa [2]. In late February, nearly 5,800 out of 6,300 US Agency for International Development (USAID) awards were cancelled, including most of those to South African organisations supporting the HIV programme [3].

## METHODS

### Tools

We used a costed version of Thembisa 4.7, a HIV transmission model representing the South African HIV epidemic that is used by SAG to prioritize HIV interventions and calculate HIV budget needs, to predict the impact of completely ceasing activities currently funded by PEPFAR on the South African HIV epidemic and the costs incurred if SAG were to take over these services [4]. For this, we calculated the incremental total costs of the public-sector HIV programme between 2025 and 2044 compared to a baseline of the current HIV programme (with a projected ART coverage of 80% of PLHIV by 2025).

Costs were estimated from the perspective of the provider, the South African public health system, in 2024 South African Rand, and converted to US dollar using the 2024 period average conversion rate of 18.33 ZAR = 1 USD [5]. Cost inputs used published data or our analyses adjusted for current SAG prices, salaries and implementation models [6], even if replacing PEPFAR-funded staff and commodities.

### Scenarios

We calculated the impact of ceasing PEPFAR-funded activities as the additional HIV infections and AIDS-related deaths between 2025 and 2044 over a baseline in which 2024 HIV programme coverage levels, including ART initiation rates, continued in perpetuity. To reflect uncertainty regarding the level and duration of reduced service coverage resulting from a cessation of PEPFAR funding, we examined four scenarios (Table 1). Our minimum scenario assumed that coverage by programme area is reduced proportionally by the percentage that PEPFAR funded in 2023, without the South African government taking over current PEPFAR activities. Our maximum scenario assumed additional impacts on the health system, e.g. through the overwhelming of existing clinics or systemic impacts of the discontinuation of PEPFAR’s support for supply chain logistics and management systems assistance. Under both scenarios, we assumed that four years of no PEPFAR funding (2025-2028) would be followed by either a recovery of programme funding over 2029-2031 to 2024 funding levels, or by no recovery reflecting lower programme funding in perpetuity. All scenarios were informed by discussions with major stakeholders in the National Department of Health.

**Table 1:** Assumed reductions in coverage by intervention and scenario from coverage levels in the 2024 HIV programme.

|  | Minimum scenario | Maximum scenario |
| --- | --- | --- |
| **ART** | 14%<sup>1</sup> | 28%<sup>2</sup> |
| **HTS** | 6% | 30% |
| **VMMC** | 45% | 70% |
| **Oral PrEP to** |  |  |
| MSM | 20% | 70% |
| FSW | 20% | 70% |
| Other adults (15+) | 0% | 30% |

*Abbreviations:* ART: antiretroviral treatment; HTS: HIV testing services; PrEP: pre-exposure prophylaxis; MSM: gay and bisexual men and other men who have sex with men; FSW: female sex workers

Finally, we estimated the cost-effectiveness of preserving each of the main four interventions with some degree of PEPFAR funding—ART, HIV testing services (HTS), oral PrEP and VMMC—under a reduced budget, by comparing each option to a baseline of a 21% defunded HIV programme, representative of the minimum scenario. We assumed that condoms, post-exposure prophylaxis, and inpatient care for people living with HIV would continue to be provided at current levels in all scenarios, as these are fully funded by SAG at baseline.

## RESULTS

A complete cessation of PEPFAR-funded activities without take-over by SAG would increase new HIV infections in adults and children over the next four years (2025-2028) by 29% to 677,158 infections in the minimum scenario, and by 56% to 822,797 infections in the maximum scenario (Figure 1A).

**Figure 1:**
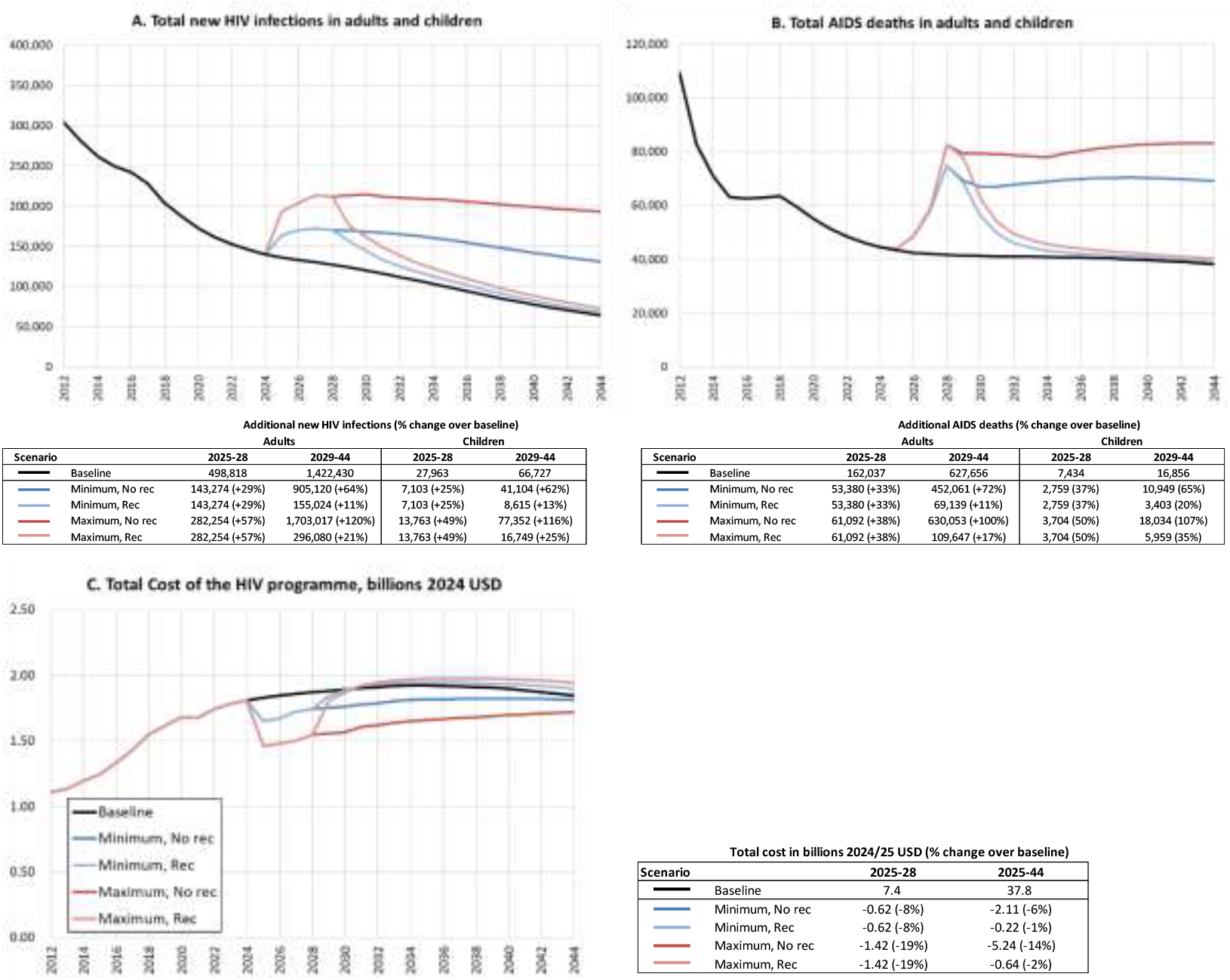
Impact of PEPFAR exit scenarios on A. new HIV infections, B. AIDS deaths and C. HIV programme costs in 2024-2044.

Over 20 years (2025-2044), new infections in adults and children would increase by 314,017 (+16%) in the minimum scenario with recovery, by 1,096,602 (+54%) in the minimum scenario without recovery, and by 608,846 (+30%) and 2,076,386 (+103%) in the maximum scenario with and without recovery, respectively (Figure 1A).

Over the next four years, AIDS-related deaths among adults would increase by 53,380 (+33%) in the minimum scenario, and by 61,092 (+38%) in the maximum scenario in adults (Figure 1B), and by 2,759 (37%) and 3,704 (50%), respectively, in children under 15 years (Figure 1B). Over the next 20 years, AIDS-related deaths in adults would increase by 122,519 (16%) in the minimum scenario with recovery, 505,44 (64%) in the minimum scenario without recovery, and by 170,739 (22%) and 691,145 (88%) in the maximum scenario with and without recovery. In children, deaths would increase by 6,163 (25%) and 13,708 (56%) in the in the minimum scenario with and without recovery, and by 9,663 (40%) and 21,738 (89%) in the maximum scenario with and without recovery (Figures 1A and 1B).

If the SAG does not fund the services and external funding does not resume (“No recovery” scenarios), total HIV programme cost would be 1-3% higher between 2029 and 2044 due to a higher number of people requiring ART and HIV inpatient services, but total cost over the full twenty years, 2025-2044, would be between 1% and 14% lower due to lower coverage of ART (Figure 1C). If the SAG took over funding of these services, the above-mentioned additional HIV infections and AIDS deaths could be averted at an estimated net additional cost over the 2025-2028 period of $622 million under the Minimum scenarios (or $1,424 million for the Maximum scenarios).

In this situation of a reduced budget, the most cost-effective option across time periods and cost-effectiveness metrics was to allocate additional funds to preserve ART and PrEP, while reducing HTS and MMC coverage equivalent to PEPFAR’s previous contribution to their expenditure (Supplementary Table 2A and 2B). When considering cost and outcomes over five years only (2025-2029), preserving ART was the most cost-effective based on both cost per death averted or cost per life-year saved, while PrEP was the most cost-effective option based on cost per infection averted (Supplementary Table 2A). Over twenty years (2025-2044), retaining PrEP alone became cost-saving over a baseline of a fully defunded HIV programme but remained the least effective at averting AIDS-related deaths (Supplementary Table 2B). The next cost-effective option, saving millions of life-years, was to maintain ART at current coverage. The option of keeping MMC alone was the least cost-effective option based on either cost per death averted or cost per life-year saved-although based on cost per infection averted, it was more cost-effective than preserving ART.

## DISCUSSION

Over the term of the current US administration, 2025-2028, the cessation of PEPFAR funding in South Africa without replacement of services by the South African government would increase expected new HIV infections in adults between 29-57% and AIDS-related deaths by 33-38%, returning to levels similar to those estimated in 2017 (for HIV infections) and 2013 (for AIDS deaths) and thereby undoing more than a decade of progress towards ending AIDS as a public health threat. Both deaths and infections will increase more sharply in children under 15 years. If affected interventions do not return to current coverage levels after 2029, either through a return in outside funding or the national government stepping in, then over 20 years there would be 1-2 million more new HIV infections and 500,000-700,000 additional AIDS deaths compared to those currently projected. Substituting these services will require additional SAG funding of between $710 million and $1.5 billion between 2025 and 2028, with the majority of these funds required to replenish antiretroviral treatment, including personnel and support services. Under a reduced budget, the most cost-effective option is to first allocate available additional domestic funds towards preserving current levels of ART.

Two previous studies have quantified the impact of discontinuing PEPFAR funding in South Africa. Assuming a halving or complete withdrawal of PEPFAR funding, Gandhi *et al*. estimated that HIV infections would increase by 286,000 and 565,000, respectively, over the next ten years [7]. Ten Brink *et al*. analysed the impact of PEPAR and European foreign assistance cuts on 26 countries with high HIV prevalence [8], estimating between 180 and 492,000 additional deaths in 2025-2030 for South Africa alone. Both estimate sets are similar to our results.

Our findings stand beside a number of limitations. Our assumptions regarding service coverage impact of discontinued PEPFAR support are based on the percentage of funding for each service provided by PEPFAR, which might not be a good proxy for the clients per service covered by PEPFAR, especially if directly PEPFAR-supported services are more expensive per person covered than government-rendered services. This could overestimate both epidemiological and budget impact, despite assuming government prices and salaries for our cost calculations. Additionally, our minimum scenario was specifically designed to represent the additional cost to SAG to compensate for discontinued PEPFAR funding, and the epidemiological consequences of this transition failing. In reality, government will partially replace services, and actual salaries and prices might be closer to those paid by PEPFAR, so actual costs and outcomes likely fall between the two scenarios. Lastly, PEPFAR-supported service discontinuation may differentially affect sub-population groups, with most key population clinics fully supported by PEPFAR. Except for PrEP, for which we stratified impacts by recipient population, our estimates applied to populations on average and might again over- or underestimate overall impact.

## Conclusion

The exit of PEPFAR from South Africa, with no replacement funding, would have devastating impacts and threaten to undo a decade of progress towards ending AIDS as a public health threat. This can be avoided if PEPFAR-funded services are taken over swiftly and comprehensively by other funders, including SAG, international donors, and private sector companies. If additional budget is limited, maintaining current ART coverage should be prioritised.

## Data Availability

All data produced in the present study are available upon reasonable request to the authors.

## ACKNOWLEDGEMENTS

GMR conceived the analysis, reviewed results and wrote the first draft. LJ performed all modelling and contributed to the first draft. EM sourced data inputs and commented on scenarios. JWIE commented on scenarios and reviewed results. LFJ developed the underlying model. All authors contributed to drafting the manuscript.

---

**Supplementary Table 1:** Expenditure on South African HIV programme in 2023 by funder [1].

| Intervention | Total amounts (2023 USD) |  |  | Percentage of total |  |  |  |
| --- | --- | --- | --- | --- | --- | --- | --- |
|  | SAG | PEPFAR | Global Fund | Total | DOH | PEPFAR | GF |
| **Treatment, care and support** | **\$1,130,133,344** | **\$192,378,465** | **\$20,328,191** | **\$1,342,840,000** | **84%** | **14%** | **2%** |
| Antiretroviral treatment | \$456,610,142 | - | \$20,050,603 | **\$476,660,745** | 96% | - | 4% |
| HIV-related laboratory monitoring | \$340,414,787 | \$623,878 | \$8,959 | **\$341,047,624** | 100% | - | - |
| Opportunistic infections (OI) prophylaxis and treatment | \$38,295 | - | \$268,629 | **\$306,924** | 12% | - | 88% |
| Programmatic activities for treatment, care and support | \$298,150,385 | \$191,754,587 | - | **\$489,904,972** | 61% | 39% | - |
| Support and retention | \$34,919,735 | - | - | **\$34,919,735** | 100% | - | - |
| Palliative care | \$539,607 | - | - | **\$539,607** | 100% | - | - |
| **Testing** | **\$52,942,909** | **\$3,393,131** | - | **\$56,336,040** | **94%** | **6%** | - |
| HIV testing and counselling | \$52,942,909 | \$3,393,131 | - | **\$56,336,040** | 94% | 6% | - |
| **Prevention** | **\$62,192,180** | **\$68,330,843** | **\$6,154,270** | **\$136,677,291** | **46%** | **50%** | **5%** |
| MMC | \$32,513,708 | \$26,822,843 | - | **\$59,336,551** | 55% | 45% | - |
| Condoms | \$19,711,908 | - | \$2,263 | **\$19,714,171** | 100% | - | 0,01% |
| PMTCT | \$6,117,827 | - | - | **\$6,117,827** | 100% | - | - |
| Prevention and promotion of testing and linkage for KP | \$607,077 | \$6,326,613 | \$2,537,434 | \$9,471,123 | 6% | 67% | 27% |
| PEP | \$937,335 | - | - | **\$937,335** | 100% | - | - |
| PrEP (AGYW) | - | \$15,496,310 | - | **\$15,496,310** | - | 100%<sup>3</sup> | - |
| PrEP (general population) | \$265,712 | \$494,740 | \$148,563 | **\$909,015** | 29% | 54% | 16% |
| PrEP (MSM) | - | \$773,571 | - | **\$773,571** | - | 100% | - |
| PrEP (prisoners) | - | \$389,148 | - | **\$389,148** | - | 100% | - |
| PrEP (SW) | - | \$196,157 | - | **\$196,157** | - | 100% | - |
| PrEP (PWID) | - | \$49,901 | - | **\$49,901** | - | 100% | - |
| Other prevention | \$1,499,006 | \$17,781,560 | \$3,466,010 | **\$22,746,575** | 7% | 78% | 15% |
| **Other programmes** | **\$925,526** | **\$-** | **\$2,744,474** | **\$3,670,000** | **25%** | **-** | **75%** |
| Social and behaviour change (SBC) programmes | - | - | \$2,744,474 | **\$2,744,474** | - | - | 100% |
| Workplace programmes | \$925,526 | - | - | **\$925,526** | 100% | - | - |
| **Total** | **\$1,250,594,095** | **\$272,764,187** | **\$35,888,383** | **\$1,559,246,664** | **80%** | **17%** | **2%** |
| System strengthening and enablers | \$170,059,771 | \$115,233,016 | \$18,377,691 | \$303,670,478 | 56% | 38% | 6% |
| **Overall total** | **\$1,420,653,866** | **\$387,997,202** | **\$54,266,074** | **\$1,862,917,142** | **76%** | **21%** | **3%** |

<sup>3</sup> Note that even though PEPFAR funded 100% of PrEP services for specific population groups in 2023, the last year that complete expenditure data has been reported for, this changed to 20% in 2024, with the SAG taking over the majority of PrEP provision, according to SAG data. In our minimum scenario, we therefore assume a reduction by 20% in PrEP coverage in these population groups.

**Supplementary Table 2A:** Cost effectiveness of preserving interventions over baseline of defunding HIV programme by 21% (2025-2029)

| Scenario | HIV infections averted | AIDS deaths averted | Life years saved | Additional cost [mil' 2024 USD] | Cost (USD)/HIV infection averted | Cost (USD)/death averted | Cost (USD)/life year saved |
| --- | --- | --- | --- | --- | --- | --- | --- |
| **Keep: ART at current coverage**<br>Reduce: HTS by 6%, MMC by 45%, PrEP in KPs by 20% | 144,816 | 55,721 | 1,274,912 | 542 | 3,740 | 9,719 | 425 |
| **Keep: HTS at current coverage**<br>Reduce: ART by 14%, MMC by 45%, PrEP in KPs by 20% | 2,162 | 331 | 7,722 | 24 | 11,271 | 73,611 | 3,155 |
| **Keep: PrEP at current coverage**<br>Reduce: ART by 14%, HTS by 6%, MMC by 45% | 987 | 2 | 56 | 2 | 1,992 | 965,563 | 35,293 |
| **Keep: MMC at current coverage**<br>Reduce: ART by 14%, HTS by 6%, PrEP in KPs by 20% | 1,958 | 4 | 104 | 49 | 25,021 | 12,501,979 | 469,177 |

**Supplementary Table 2B:** Cost effectiveness of preserving interventions over baseline of defunding HIV programme by 21% (2025-2044)

| Scenario | HIV infections averted | AIDS deaths averted | Life years saved | Additional cost [mil' 2024 USD] | Cost (USD)/HIV infection averted | Cost (USD)/death averted | Cost (USD)/life year saved |
| --- | --- | --- | --- | --- | --- | --- | --- |
| **Keep: PrEP at current coverage**<br>Reduce: ART by 14%, HTS by 6%, MMC by 45% | 1,144 | 158 | 3,937 | -6 | Cost-saving |  |  |
| **Keep: ART at current coverage**<br>Reduce: HTS by 6%, MMC by 45%, PrEP in KPs by 20% | 266,562 | 125,047 | 2,899,090 | 187 | 703 | 1,499 | 65 |
| **Keep: HTS at current coverage**<br>Reduce: ART by 14%, MMC by 45%, PrEP in KPs by 20% | 7,765 | 2,146 | 50,810 | 7 | 918 | 3,322 | 140 |
| **Keep: MMC at current coverage**<br>Reduce: ART by 14%, HTS by 6%, PrEP in KPs by 20% | 38,686 | 1,499 | 38,216 | 27 | 698 | 18,007 | 706 |

## Footnotes

1 This means a resulting 86% of baseline ART coverage (82%), i.e. 71% of PLHIV being covered.

2 This means a resulting 72% of baseline ART coverage, i.e. 60% of PLHIV being covered.

